# Genomic epidemiology of SARS-CoV-2 infections in The Gambia, March 2020 to Jan 2022

**DOI:** 10.1101/2022.09.07.22278739

**Authors:** Abdoulie Kanteh, Haruna S. Jallow, Jarra Manneh, Bakary Sanyang, Mariama A. Kujabi, Sainabou Laye Ndure, Sheikh Jarju, Alhagie Papa Sey, Dabiri K Damilare, Yaya Bah, Sana Sambou, Gibril Jarju, Buba Manjang, Abubacarr Jagne, Sheikh Omar Bittaye, Mustapha Bittaye, Karen Forrest, Desta Alamerew Tiruneh, Ahmadou Lamin Samateh, Sheriffo Jange, Stéphane Hué, Nuredin Muhammed, Alfred Amambua-Ngwa, Beate Kampmann, Umberto D’Alessandro, Thushan I. de Silva, Anna Roca, Abdul Karim Sesay

## Abstract

**Background:** COVID-19, caused by SARS-CoV-2, is one of the deadliest pandemics over the last 100 years. Sequencing is playing an important role in monitoring the evolution of the virus, including the detection of new viral variants. This study describes the genomic epidemiology of SARS-CoV-2 infections in The Gambia.

**Methods:** Nasopharyngeal and/or oropharyngeal swabs collected from suspected cases and travellers were tested for SARS-CoV-2 using standard RT-PCR methods. SARS-CoV-2 positive samples were sequenced following standard library preparation and sequencing protocols. Bioinformatic analysis was done using ARTIC pipelines and lineages assigned using Pangolin.

**Findings:** Between March 2020 to January 2022, there were almost 12,000 SARS-CoV-2 confirmed cases distributed into four waves, each of them lasting between 4 weeks and 4 months, with more cases during the rainy seasons (July-October). As shown by the 1643 sequenced samples, each wave occurred after new viral variants and/or lineages were introduced in The Gambia, generally those already established in Europe and/or in other African countries. Local transmission was higher during the first and third wave, with mostly B.1.416/Senegal/Gambian lineage and AY.34.1/Delta subtype, respectively. The second wave was driven by two variants, namely Alpha and Eta and B.1.1.420 lineage. The Omicron/fourth wave was the shortest.

**Interpretation:** Efficient surveillance, including strengthening entry points and screening asymptomatic individuals especially during the rainy seasons would be important to promptly detect and control future waves in The Gambia and the subregion.

**Funding:** Medical Research Unit The Gambia at LSHTM, UK Research and Innovation funding (grant reference MC_PC_19084), MRC/UKRI MC_PC_19084 and World Health Organisation.

## Introduction

The COVID-19 pandemic caused by the severe acute respiratory syndrome coronavirus 2 (SARS-CoV-2) has claimed more than six million lives and infected over 526 million people globally [Accessed: 15-03-2022]. ^1^ As of May 2022, Africa has contributed to 2·0% of cases and 2·7% deaths to the global COVID-19 burden, ^2^ despite constituting 17% of the world’s population.^3^ In contrast, Europe, with more than two million SARS-CoV-2 infections, contributed to 42% of cases and 32% of deaths. ^2^ The pandemic has evolved differently around the globe starting later in Africa than in other regions ^4^ although sero-prevalence studies suggest that transmission has been much higher than reported. ^5–7^

Despite its low mutation rate, widespread transmission and high case numbers have increased the diversity of SARS-CoV-2 during the pandemic.^8^ Prolonged infections in immunocompromised individuals ^9^ and high rate of reinfections increase the likelihood of new mutations, leading to the emergence of new variants. More importantly, introduction of new variants of concerns (VOCs) reported in different countries ^10,11^ is responsible for higher transmission, ^12^ disease severity ^13,14^ and immune evasion ^15^. For example, the Alpha variant or B·1·1·7 lineage, first identified in the UK in November 2020, was reported to be between 43% and 90% more transmissible than the wild-type SARS-CoV-2. ^16^ The Delta variant/B·1·617·2, first reported in India in October 2020, is 4 times more likely to evade the immune system and 50 times more transmissible than the Alpha variant. ^17^ Indeed, the emergence and spread of the Delta variant changed the dynamic of the epidemic in many African countries as it coincided with an increased numbers of reported cases and deaths, with a very long and slow downward trend. ^18^ The Omicron variant, first identified in South Africa in November 2021, was described as the most transmissible variant, quickly spreading across the world and exhibiting a high degree of evading antibody immunity.^19^

Effective strategies to quickly detect new variants, sources of infections, outbreaks and transmission patterns in different geographical settings are essential for controlling the spread of SARS-CoV-2. Whole genome sequencing enabled detailed understanding of pathogen’s evolution and of its population structure. ^20^ High resolution viral genome sequences linked with epidemiological data allowed identifying the origin of the virus ^21^, and monitoring its spread and evolution, both globally ^22^ and locally. ^23,24^ Moreover, the information obtained from sequencing is essential for guiding the design of the next generation vaccines. ^25^ As of May 2022, more than 11 million genomes have been sequenced globally and submitted to GISAID. ^26^ Unfortunately, only 1% of these sequences is from African countries ^26^, a major obstacle to understanding the local evolution of the virus.

The Gambia, a small country in West Africa, identified its first imported COVID-19 case from the UK ^27^ in March 2020. By February, 2022, The Gambia had recorded almost 12,000 cases and 365 deaths.^28^. More than 1,600 SARS-CoV-2 genomes have already been sequenced and analysed with the aim of providing real time genomic data to monitor circulating variants within The Gambia. Moreover, this information is essential for monitoring local transmission and will significantly contribute to the global genomic dataset. We have already described the origin of the first five cases diagnosed in The Gambia during the early phase of the pandemic ^27^ and used phylogenetic analysis to confirm SARS-CoV-2 reinfection in two healthy individuals. ^29^ The study presented here aims to extensively describe the genomic epidemiology of the first four SARS-CoV-2 waves observed in The Gambia over a 23-month study period.

## Methods

### The Gambia, population demographics and climate

The Gambia has a population of about 2·5 million people, predominantly Muslims, with a median age of 17·8 years. ^30^ More than half of the population live in urban areas, mainly at the coast. The climate is subtropical with two seasons, i.e., a rainy season between June and October and a dry season for the rest of the year, although November and December are “wet months” during which the malaria transmission peak occurs. ^31^ Average temperatures range between 23°C and 33°C during the rainy season with high humidity (>80%) and between 18°C and 30°C during the dry season. ^31^

### The Gambia Healthcare system

The Gambian Government is the main healthcare provider, operating seven referral hospitals, eight major health centres and 16 minor ones across the country. Healthcare delivery is mostly through the primary or local health posts. In addition, there are about 30 private and nongovernmental organisation clinics across the country. ^32^

### COVID-19 response in The Gambia

Measures implemented to control the spread of the COVID-19 epidemic have already been described elsewhere. ^33^ Briefly, international borders were closed in March 2020 and a state of emergency declared (Figure 1). Initial SARS-CoV-2 testing by PCR was focused on identifying imported cases and tracing, plus isolating case contacts, especially among travellers from Senegal and their contacts.

**Figure 1.**
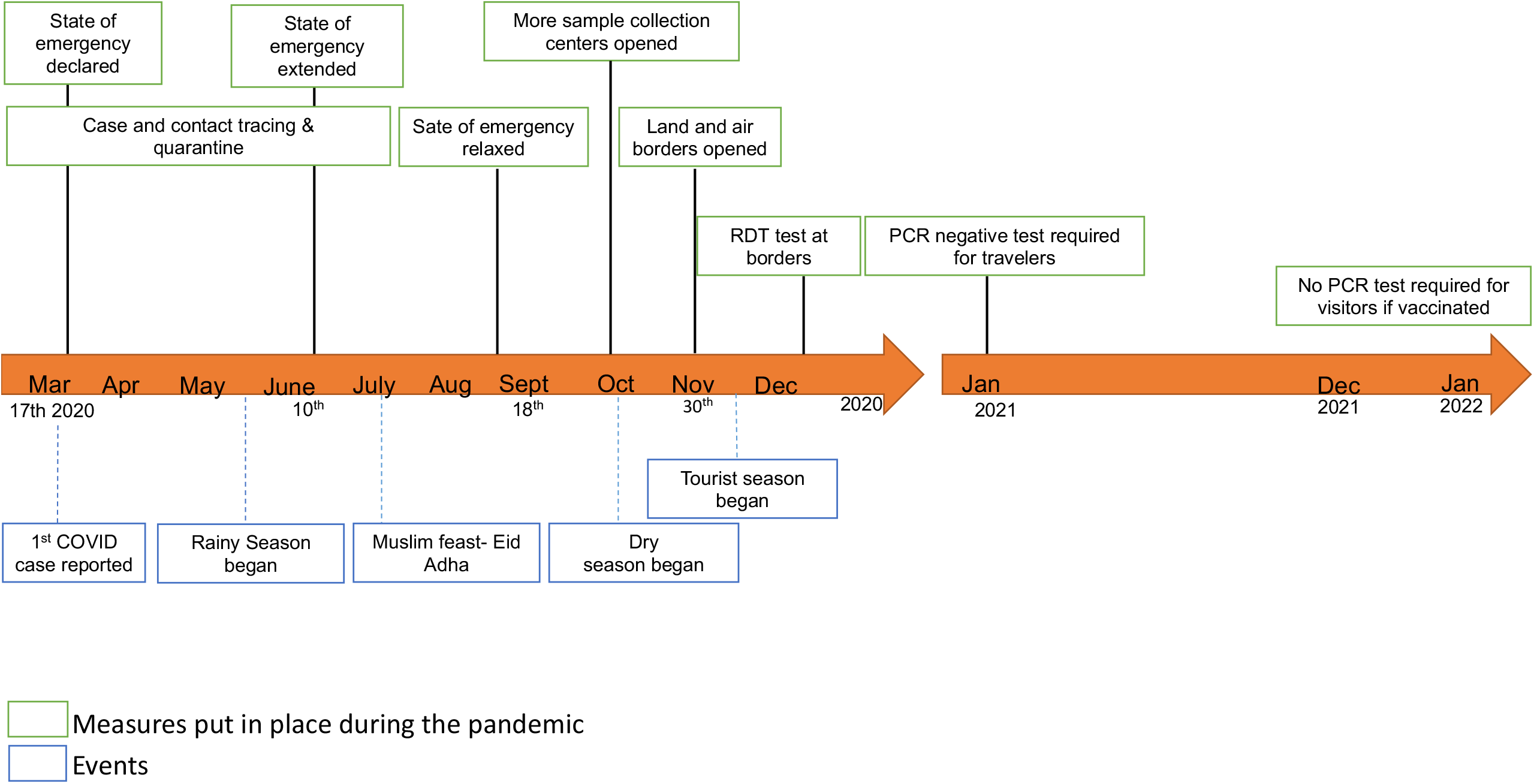
Timeline of events and measures put in place during the pandemic.

### Sample collection

Nasopharyngeal, oropharyngeal or both swabs were collected for SARS-CoV-2 detection using FLOQSwabs. ^34^ Although there were different reasons for testing, most samples (>85%) were collected as a pre-travel testing requirement (FigureS1).^35^ Swabs were placed in single tubes containing a universal transport medium. ^34^ and delivered to the laboratory within 24 hours. Demographic data including gender, age, address, travel history and any other relevant information were collected whenever possible.

**Figure S1**. Distribution of sample collection (reason for testing) in The Gambia over the 23 months of study period.

### RNA extraction and PCR

Samples were transported for analysis to the National Public Health Laboratory (NPHL) or the MRC Unit The Gambia at LSHTM molecular diagnostic laboratory. Sample processing, RNA extraction and real-time reverse transcription PCR (RT-PCR) were done following standard WHO guidelines.^33,36^ From August 2020 to date, PCR positive SARS-CoV-2 required PCR amplification of both the screening (E) and confirmatory (N) genes. Results and associated metadata collected for each sample were recorded in RedCap ^37^ and DHIS2 ^38^, respectively.

### Sample selection profile

Samples without basic metadata such as date of collection, age and sex were not processed (Figure S2). Criteria for sequencing changed over time. From March to June 2020, all samples were processed and sequenced, regardless of cycle threshold (Ct) values. However, due to the high proportion of failed sequencing among samples with Ct values above 30, in June 2020 it was decided to sequence only samples with a Ct value of ≤ 30. In addition, samples with low DNA concentration ((≤ 4ng/μl) after library preparation were also excluded.

**Figure S2**. Total count of samples that were dropped from the study and reason for dropping.

### Library preparation and sequencing

cDNA conversion and multiplex PCR were prepared following ARTIC nCoV-2019 protocol. ^39^ Updated primer schemes were always used to ensure optimum sequencing of emerging VOCs. ^39–42^ Due to variable Ct values, all RNA samples were run for 35 cycles. Pooled PCR products were either cleaned up using 1x SPRI/AMPure XP beads or library prepared directly without bead clean-up following manufacturer’s instructions. Libraries were prepared for sequencing on the Oxford Nanopore platform as described previously. ^44^

### Quality Control

Samples were prepared and sequenced in 96-well plates with one cDNA and RNA extraction negative control each, per plate. Consensus sequence was defined as passing quality control if greater than 50% of the genome was covered by confident calls or there was at least one contiguous sequence of more than 10,000 bases and no evidence of contamination in the negative control. A confident call was defined as having 10x depth of coverage. If the coverage fell below these thresholds, the bases were masked with the character N, indicating that the base at that position is unknown or not available. Low quality SNP were also masked with Ns. For phylogenetic analysis, only genomes with less than 10% missing bases were used to construct robust phylogenetic trees.

### Clustering and phylogenetic analyses

Raw reads were analysed using the ARTIC bioinformatics pipeline.^45^ FASTQ files were trimmed to remove adapters and mapped to a reference genome (GenBank accession number MN908947·3). Consensus genomes were further analysed to assigned lineages and clades using the Pangolin online tool ^46^ and Nextclade, ^47^ respectively. To find the closest global sequences to that of Gambian samples from GISAID database, we first stratified our sequences into different waves (wave 1 to 4). We then used Uvaia (GitHub, last accessed: 14-05-2022) to find the closest sequences to our samples against from global sequences deposited in GISAID [accessed 14-05-2022]. Only sequences (genomes in GISAID) with high nucleotide similarity (raked as 1 in Uvaia) to Gambian samples were retained for phylogenetic analysis. Ambiguous bases and low coverage regions were masked prior to multiple sequence alignment using MAFFT (v7·453).^48^ Maximum likelihood phylogenetic trees were constructed using IQTREE (v1·3·11·1) ^49^ under the HKY model of nucleotide substitution, and branch supports were estimated using ultra-fast bootstrapping, as implemented in IQTREE, with 1,000 replicates. The phylogenies were visualised and edited using microreact. ^50^

## Results

### Samples, cases, and sequences

Between March 2020 and January 2022, among the 75,554 samples tested for SARS-CoV-2, 15·8% (11,911) were positive [Accessed:13-02-2022].^2^ Among the 3,254 samples received for sequencing, 1643 (50.5%) passed all QCs and were included in this study (Figure 2). Reasons for missing sequenced data are detailed in figure S2

**Figure 2.**
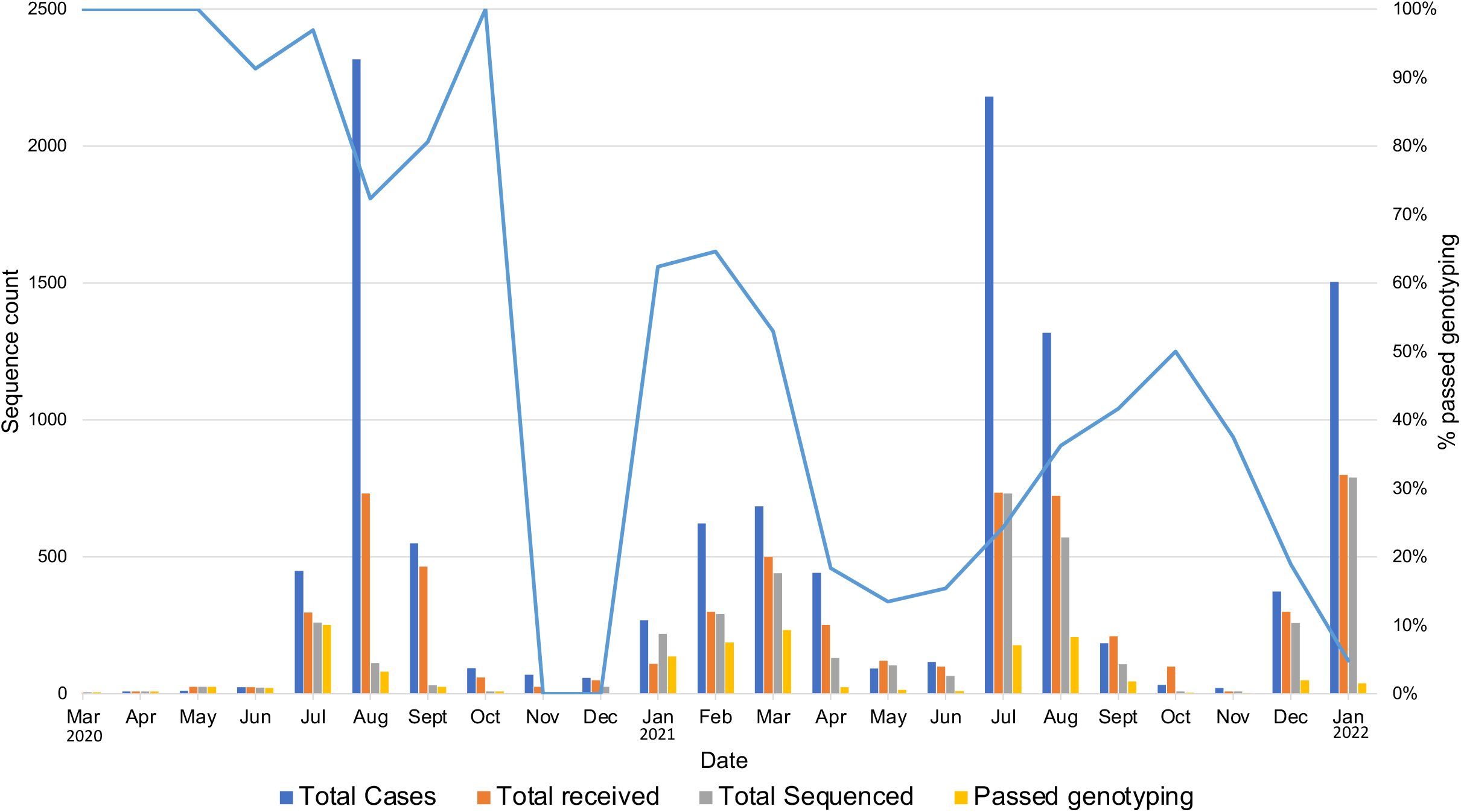
Number of SARS-CoV-2 positive samples sequenced by month. The bars represent the total number of cases (blue), total number of samples received for sequencing (orange), total samples sequenced (grey) and total passed genotyping (yellow). The blue line represents the proportion of samples sequenced that passed genotyping.

### Description of epidemic waves

The Gambia has experienced four different waves since the first identified case of SARS-CoV-2 infection (Figure 3). The first wave, predominantly associated with the B·1·416 lineage started in July 2020 and lasted for 3 months. The second wave started in January 2021 and ended in May 2021 and was milder (less cases and deaths) and lasted longer than the first wave. Variants Eta/B·1·525 and Alpha/B·1·1·7, and a lineage (B·1·1·420), dominated during the second wave. The third wave, with a higher number of cases and deaths, occurred between July and September 2021, and was driven by the Delta/B·1·617·2 variant and its subtypes. The fourth wave, between December 2021 and January 2022, was dominated by the Omicron variant/subtypes.

**Figure 3.**
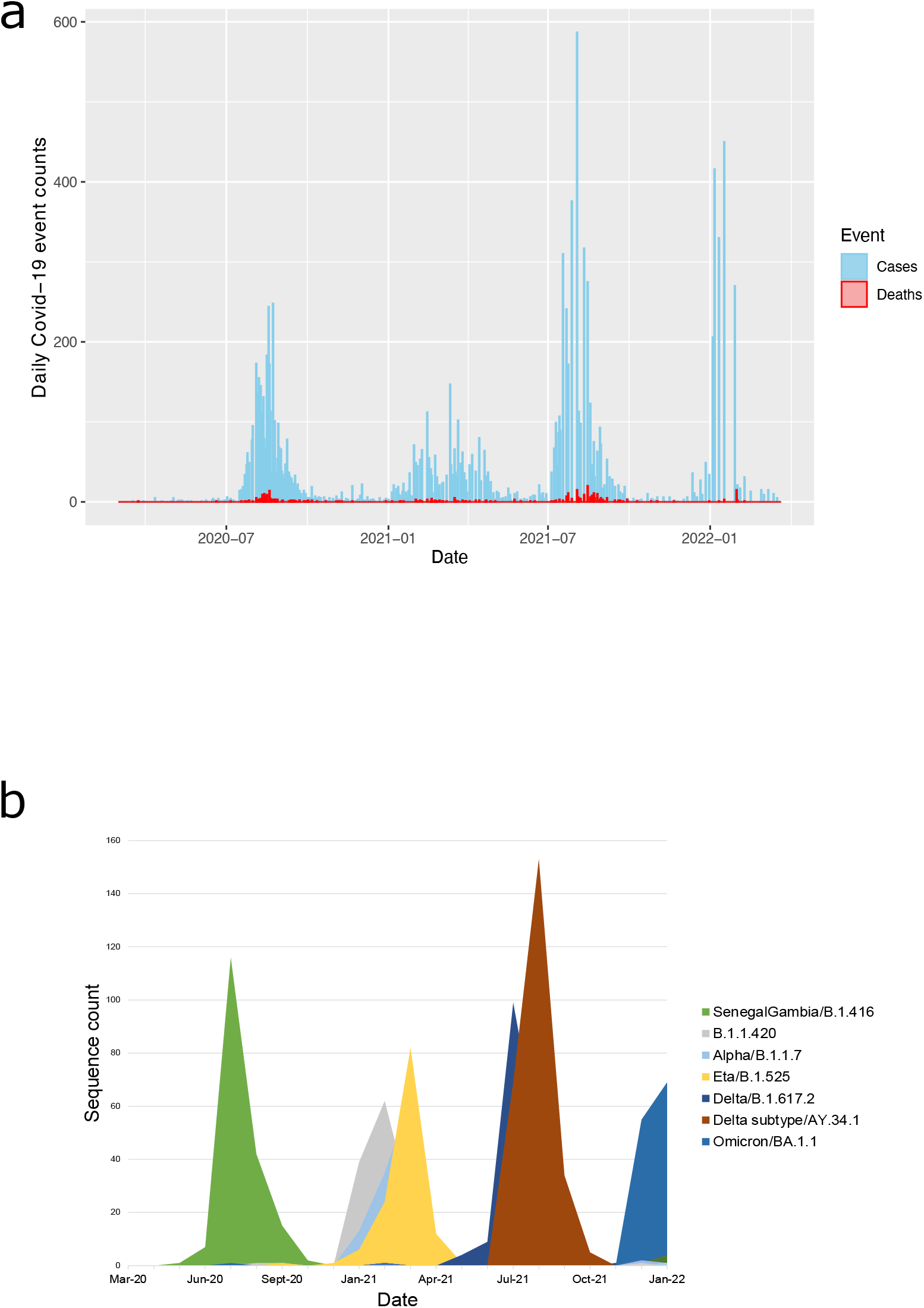
Number of reported SARS-CoV-2 overtime (a) reported cases and deaths (b) trend of lineages and major variants in The Gambia from March 2020 – January 2022.

### Changes in variant prevalence

Between March 2020 and January 2022, 55 different lineages were identified (Figure 4). During the first wave, lineage B·1·416 represented 49% (173/354) of the total samples sequenced followed by B·1·1 16% (57/354) (Figure S3a). Six lineages were observed in only one sample (B·1·525, B, B·1·177, A·27, B·1·1·7 and B·1·582).

**Figure 4.**
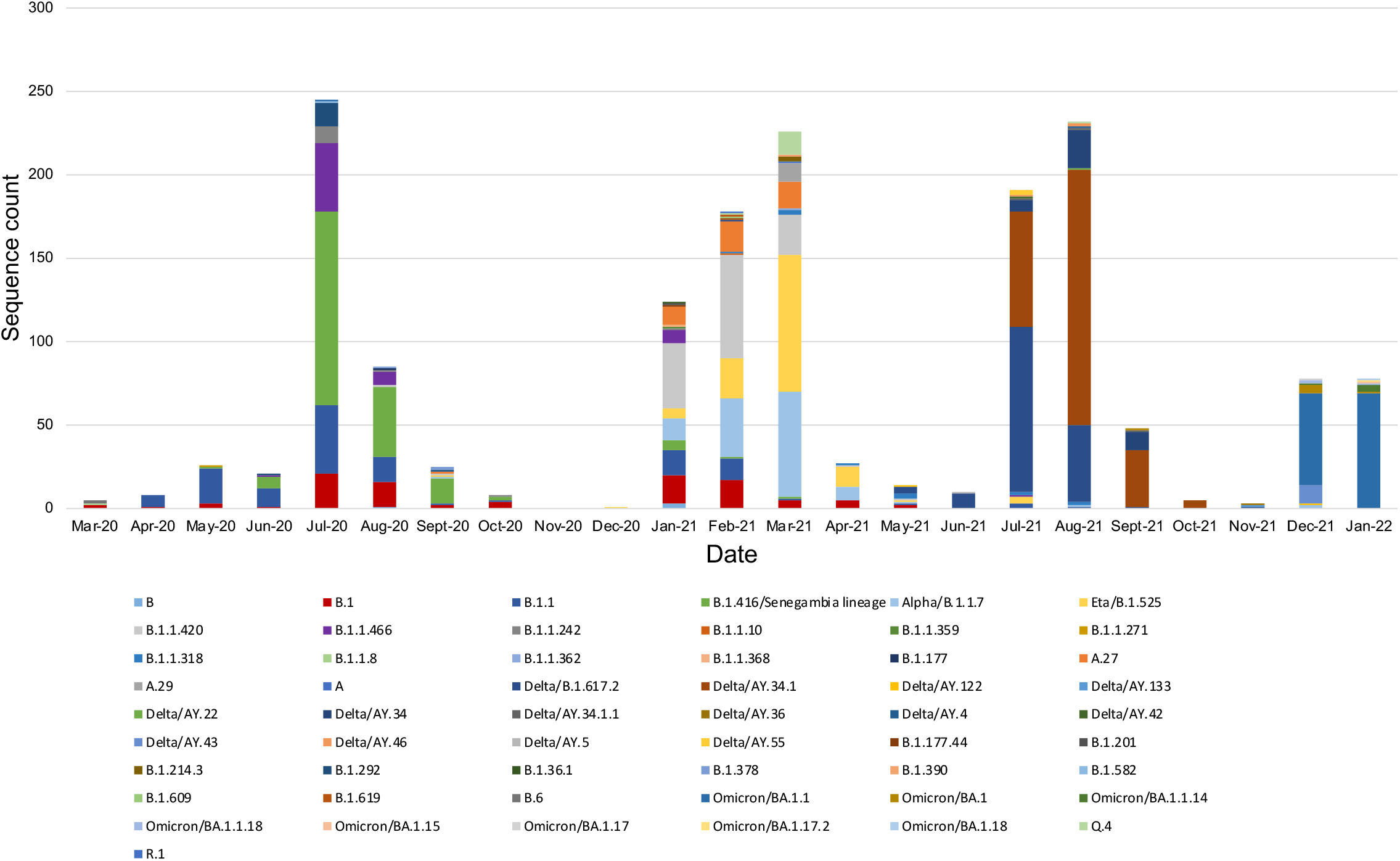
The number of different lineages identified each month during each wave from March 2020 – January 2022.

**Figure S3**. The proportion of lineages by wave during the epidemic.

Some lineages identified during the first wave disappeared during the second wave, while new lineages emerged. For example, B·1·416 represented only 1% (8/555) of lineage sequenced from the second wave, with the last identified in February 2021 (Figure S3b). In March 2021, almost one year after the identification of the first case in The Gambia, B·1·1·420 became the dominant lineage constiuting 23% (126/555) of isolates, followed by Eta/B·1·525 (22%, 124/555) and Alpha/B·1·1·7 (21%, 119/555) respectively (S3b). Fifteen lineages were represented by only one sample.

The Delta/B·1·617·2 variant was first confirmed in The Gambia in early April 2021 and became the dominant variant within a few weeks (Figure S3c). Between July and August 2021 (peak of the third wave), the Delta variant subtype AY·34·1 represented more than half (256/470) of all the samples sequenced, while the rest consisted of other lineages such as B·1·617·2 (31%, 146/470) and AY·34 (9%, 41/470), among others. Six lineages were observed in only one sample (AY·22, AY·4, AY·42, B·1·466, B·1·17 and Q·4).

The omicron variant and its subtypes dominated the fourth wave (Figure S3d). Subtype BA.1.1 accounted for 79% (124/156) of all the genomes sequenced. The remaining samples were BA·1 representing approximately 7% (6/156) and the Delta variant subtypes, representing 8% (12/156) of all samples, with subtype AY.34 being the most common. Five lineages were observed in only one sample (AY·122, BA·1·15, BA·1·17, BA·1·17·2, and BA·1·18).

### Clustering and Phylogenetic analysis

Phylogenetic analysis of Gambian samples revealed eight distinct, strongly supported clades, which clustered based on lineage types. (Figure 5). Phylogenetic clustering using closest global sequences showed high local transmission events during the first and third wave. (Figure 6a and 6c). Lineage B·1·416 clustered with genomes from Senegal, suggesting spread of infections between these two countries (Figure 6a). Similarly, high clustering among Gambian Delta samples during the third wave showed possible importation and subsequent spread via local transmission. Overall, cluster analysis showed possible importation of the virus mainly from Europe and Africa.

**Figure 5.**
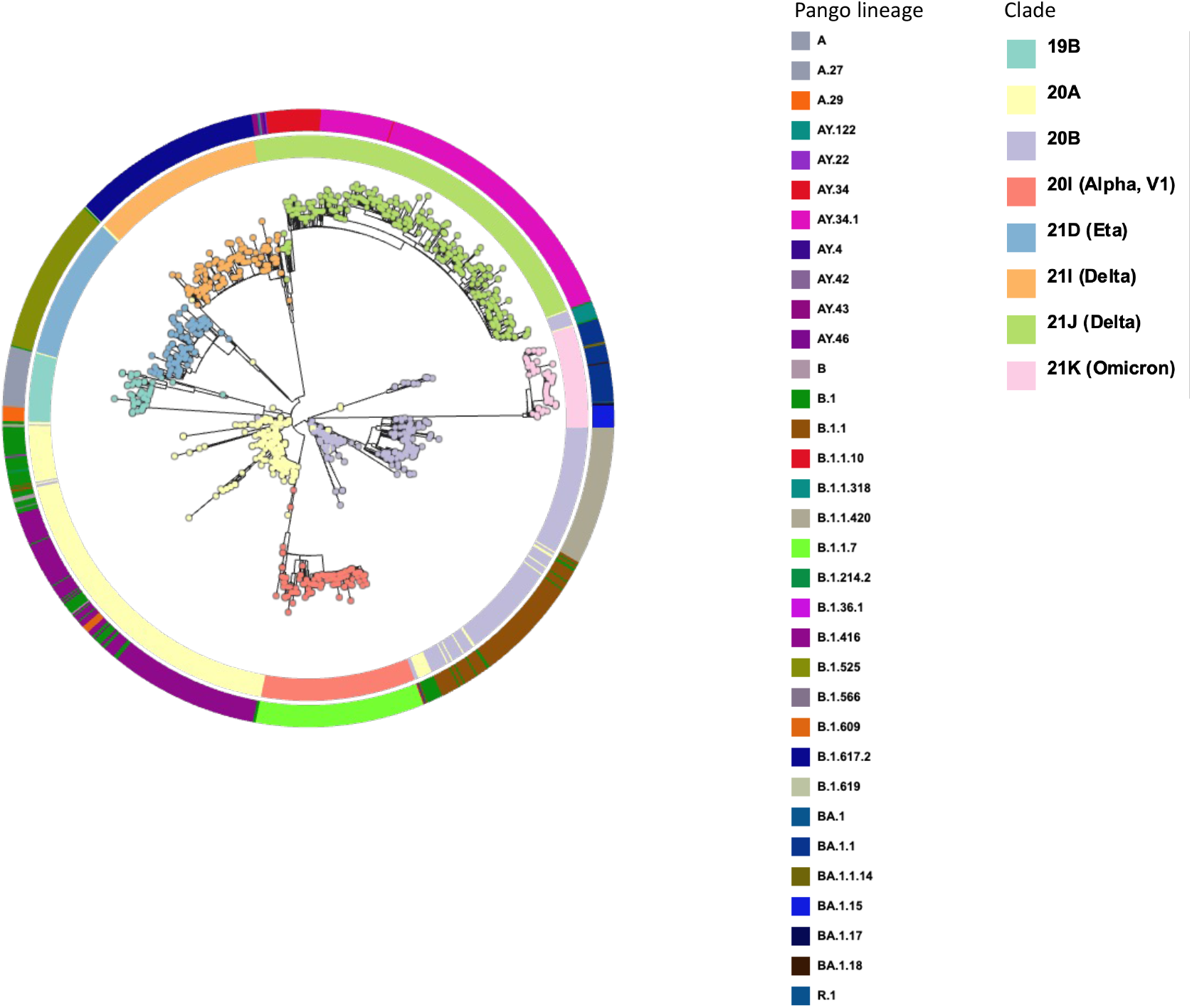
Maximum likelihood tree of 1,313 SARS-CoV-2 genomes sampled in The Gambia between March 2020 – January 2022. Clades are labelled A through H with lineages or variants associated with each clade shown.

**Figure 6.**
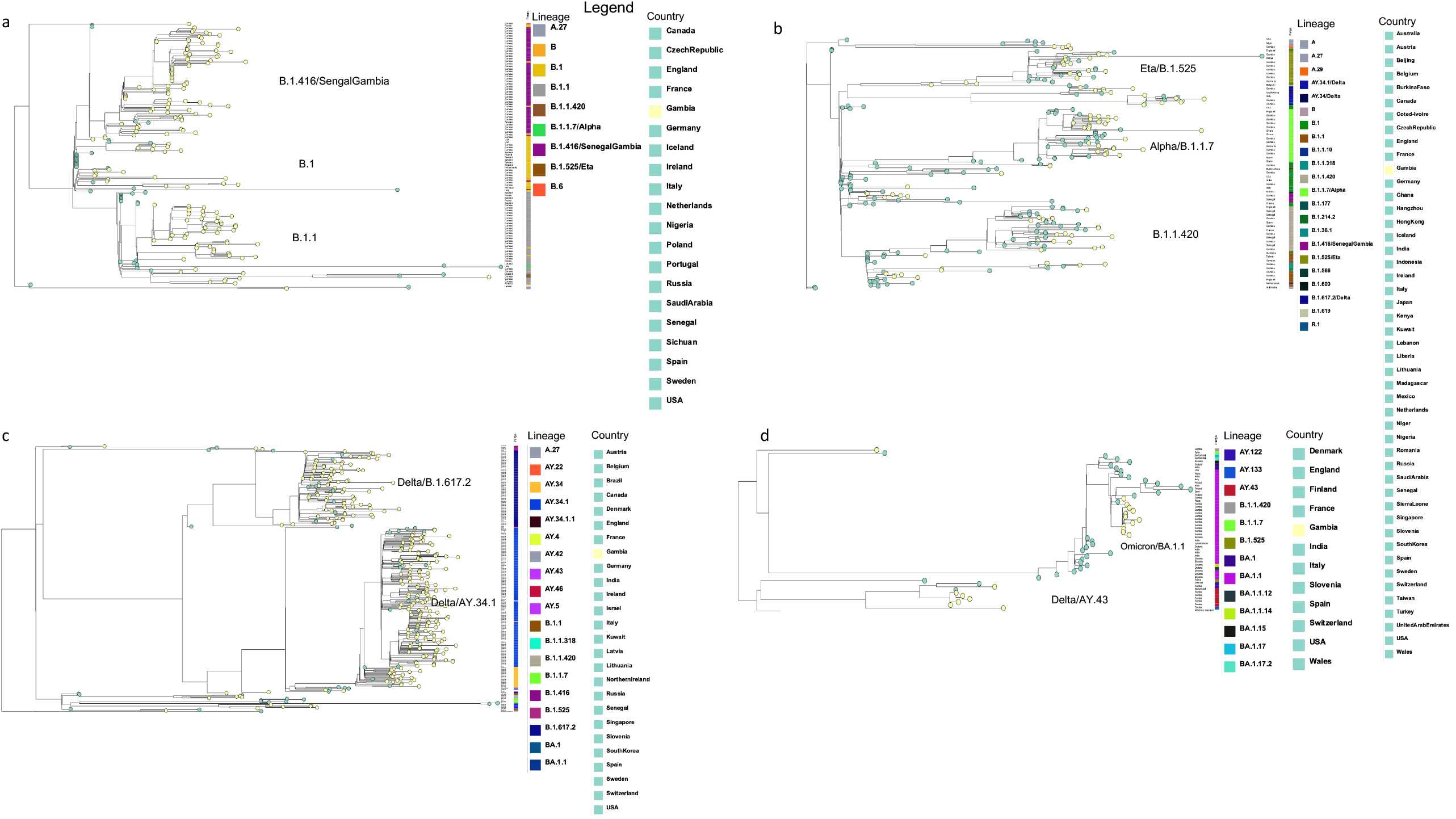
Maximum likelihood tree of the closest SARS-CoV-2 genomes to those sampled from The Gambia between March 2020 – January 2022. Each tree represents a specific wave, waves 1 - 4 (labelled a - d) with branch tips representing the country where closest global sequences were sampled from. All trees were rooted on the SARS-CoV-2 reference genome (GenBank accession number MN908947·3).

## Discussion

We have presented a comprehensive description of the genomic epidemiology of the SARS-CoV-2 epidemic in The Gambia. Following the first detected case diagnosed in March 2020, there were four different waves, two waves in the rainy season and two in the dry season, each lasting between 4 weeks and 4 months. All waves ended despite the lack of major restrictions to stop the epidemic. New waves occurred when new variants and/or lineages entered the country, generally those already prevalent in Europe, and other countries at the time. Phylogenetic clustering revealed high local transmission during the summer (first and third wave) with possibly more imported cases during the second wave.

Both the first and the third waves were short, lasting less than three months but intense. They occurred during the rainy seasons and started almost at the same time in 2020 (B·1·416 lineage) and 2021 (Delta variant), when humidity is more than 80% and maximum temperatures above 33°C. ^31^ The effect of temperature and humidity on SARS-CoV-2 tranmission is unclear, ^51,52^ but this observation is consistent with transmission dynamics described for other respiratory viral pathogens for which the rainy season coincides with the period of highest incidence.^53^ Even if the virus infectivity is not modified by climate parameters, people are more likely to spend more time indoors during heavy rains and this may increase the likelihood of household transmission. ^33^ The initial hypothesis that lower temperatures could be associated with higher SARS-CoV-2 transmission ^54^ with increased susceptibility to infection due to irritation of nasal mucosa especially in temperate regions ^55^ does not fit with the occurrence of the second wave in The Gambia. Despite the introduction of very transmissible variants, Alpha and Eta. ^56^, the epidemic peak was lower but lasted longer in this second wave when compared to the first and third waves.

In contrast with Senegal and Burkina Faso, where a sharp increase in cases was seen just after the first few confirmed cases ^33^, the first epidemic wave in The Gambia did not immediately followed the diagnosis of the first case. This is probably due to the declaration of the state of emergency closing The Gambia’s air and land entry points. Although genomic analysis from the first five cases in The Gambia confirmed importation of the virus from Europe and Asia^33^, the close genomic relatedness of Gambian sequences during the first wave with viruses sampled in Senegal suggests that importation of the new strains occurred mainly by land as air enforced travel restrictions lasted until the end of the first wave. Enforcing land border closures is also materially more difficult than air travel restrictions. These genomic observations are epidemiologically probable as the sudden increase in the number of cases in July 2020 coincided with the preparations for the major Muslim feast of Eid-al Adha, where an inflow of undetected cases through the porous Senegal-Gambia border may have occurred. This concurs with the high prevalence of the so-called Senegal/Gambia lineage or B·1·416 ^11^, which also dominated the first wave in Senegal (March – August 2020).^57^ This lineage is characterised by few mutations across the genome, the most notable being the D614G on the spike protein gene linked to increased viral load and high transmission, but not to increased disease severity.^58^ So far, The Gambia and Senegal have reported the highest number of B·1·416 cases.^26^ Other African countries such as Burkina Faso and Morocco have also reported this lineage, but only two genomes each.^26^ The low number of isolates of this lineage in other West African or neighbouring countries could be due to lack of sequencing facilities in the region, emphasising the importance of genomic sequencing for monitoring the evolution of the virus during an epidemic. Nevertheless, South Africa sequenced more genomes than any other country in Africa^26^ and did not identify this lineage. Importantly, high clustering among Gambian sequences suggests high local transmission, mostly associated with B·1·416 lineage. Consistent with our findings, importations of SARS-CoV-2 within Africa increased as more cases were reported in Africa due to high local transmission. ^59^ On the other hand, phylogenetic analysis of Gambian genomes along with global sequences also showed relatedness with samples from Europe.

The second wave in The Gambia was probably favoured by the reopening of borders and relaxation of lockdown measures in September 2020, and the influx of tourists from Europe and Africa during and immediately after Christmas. The introduction of more lineages/variants in the country and indeed the clustering of Gambian sequences with those from different parts of the world further confirms the effect of travelling/tourism in transmission.^60^ This wave was predominantly caused by two variants, Alpha, and Eta and B·1·1·420 lineage. In contrast to reports from the UK and elsewhere^16^, the Alpha variant, although more transmissible than pre-existing lineages, was associated with slightly lower number of sequenced cases during the wave compared to Eta and B·1·1·420 lineage. A similar pattern was also reported in Nigeria where Alpha and Eta co-circulated.^61^ Using mathematical modelling, Zhao and his colleagues suggested that Eta seemed slightly more infectious than Alpha in Nigeria. Phylogenetic analysis showed high clustering of most of the Alpha Gambia samples with samples from Germany, Spain, England as well as Ghana while Eta samples clustered more with UK, Germany and Belgium. The B·1·1·420 lineage, reported to originate from Senegal ^57^, dominated the second wave in both countries. Overall, sequences from The Gambia clustered closely mainly with sequences from Senegal and from Europe.

The third wave was caused by the more contagious Delta variant and its subtypes, resulting in higher number of daily cases.^62^ In The Gambia, the peak was in July-August 2021 and was associated with higher mortality, similar to other African countries, with deaths peaking during the week of 19 July 2021.^62^ According to our phylogenetic analysis, most of the Gambian Delta sequences were highly related, suggesting high local transmission. Few genomes clustered with sequences from Europe, Africa (Senegal only), Asia as well as North America suggesting possible importation from these countries. In contrast to other circulating variants, Delta evolved into two distinct clades, suggesting that the variant may have gone through positive selection due to the acquisition of many mutations. This divergence was driven by AY·34·1 sub-lineage forming a single cluster associated with more than half (54%) of the samples sequenced in The Gambia during this wave. High prevalence of this lineage was reported in Senegal during the same period, with only few seen in other parts of Africa ^26,63^, highlighting again the proximity of these two countries.

The fourth wave caused by the Omicron variant was characterised by the sharpest increase in cases to date followed by a drastic decrease in less than a month. ^64^ In South Africa, where Omicron was first detected in November 2021, the daily cases peaked in December and quickly after, declined.^65^ A similar trend was seen in many other countries, including Eastern and Central Africa, and Europe. ^65^ The rapid decline of cases may be due to the high infectivity of the virus (Omicron) leading to high percentage of infection in the population, subsequently decreasing the number of susceptible individuals. ^66^

The fourth wave coincided with the Gambian tourist season (October – February) when fully vaccinated individuals were allowed into the country, without needing a negative PCR result. This probably resulted in new introductions followed by local transmission. As expected, phylogenetic analysis showed more Gambian omicron sequences clustering with those from Europe, the origin of more than half of all tourist. ^67^ In addition, BA·1·1 subtypes, associated with high infection rates in Europe and North America was the most dominant subtype in the Gambia. The omicron variant encodes more than 30 amino acid substitutions in the spike protein, enhancing its ability to evade the immune system ^52^ and thus spreading faster and more likely to re-infect previously infected and/or vaccinated individuals. A significant number of cases may have been missed as the requirement for a negative PCR for outgoing travellers, who represent almost 90% of the overall SARS-CoV-2 cases, was waived. Interestingly, Delta variant subtypes were still circulating during the fourth wave, most notably the AY·43 sub-lineage reported in more than 130 countries and 54 US states^11^.

Our study has some limitations to be considered when interpreting the results. Systematic testing of individuals with flu-like symptoms in The Gambia has been limited throughout the pandemic and therefore most sequences were obtained from samples collected from asymptomatic outgoing travellers. There is probably a substantial under-estimation of the overall case numbers and some lineages circulating in the country may have been missed. Most samples were collected at the coast and lineages circulating exclusively upcountry may have been missed. The inference of higher transmission and severity of the third wave, mainly Delta variant, although expected as per international data may have been biased by a potential change in behaviour towards testing sick individuals (severity) or the increased number of outgoing travellers coinciding with the summer months of 2021 when international flights were open. Most of the detected cases in The Gambia were outgoing travellers. In addition, most positive samples failed our quality checks and hence were either not sequenced or failed analysis.

Our analysis of SARS-CoV-2 epidemic in The Gambia shows an annual trend, with higher transmission during the rainy season, in line with transmission patterns of other respiratory viruses. Phylogenetic analysis confirmed multiple introductions of the virus from different parts of the world, mostly from Senegal during the first wave, and subsequently from Europe and Asia. As expected, most of the lineages described were imported, later spreading locally. New waves coincided with the introduction of new lineages, highlighting the importance of implementing well-structured and active genomic surveillance at national level to detect and monitor emerging and circulating variants. In-depth analysis using phylodynamic inference could elucidate the transmission dynamics and importation patterns of the virus over the course of this and future pandemics.

## Supporting information

Ethical approval letter

## Data Availability

All data produced in the present work are contained in the manuscript

## Declaration of interest

The authors declare no conflict of interest

## Acknowledgement

The genomic COVID-19 surveillance in The Gambia is conducted by Ministry of Health (National public health lab) in collaboration with Medical Research Council Unit The Gambia at the London School of Hygiene & Tropical Medicine (MRCG at LSHTM). The authors wisht to acknowledge the support of all staff at National Public Health laboratory, Directorate of Planning and Information and Epidemiology and Disease Control. We would also thank the lab team at MRCG at LSHTM, field workers, nurses, and data management. AK wish to thank Dr Andrew Page and team at the Quadram Institute for their support. We are grateful to all participants whose samples were used in this study. Finally, we thank all laboratories that deposited their sequences in GISAID for public access.

## Notes

### Competing Interest Statement

The authors have declared no competing interest.

### Funding Statement

This study was funded by Medical Research Council Unit The Gambia at LSHTM, UK Research and Innovation funding (grant reference MC_PC19084), MRC/UKRI MC_PC_19084 and World Health Organisation.

### Author Declarations

The Gambia Government/MRCG Joint Ethics Committee Request ID/Ethics ref:L2002.E06 Request Title: Request for ethical waiver accepted

