## Supplementary figures and images for "Genomic epidemiology of SARS-CoV-2 infections in The Gambia, March 2020 to Jan 2022"

### Supplemental Figure 1

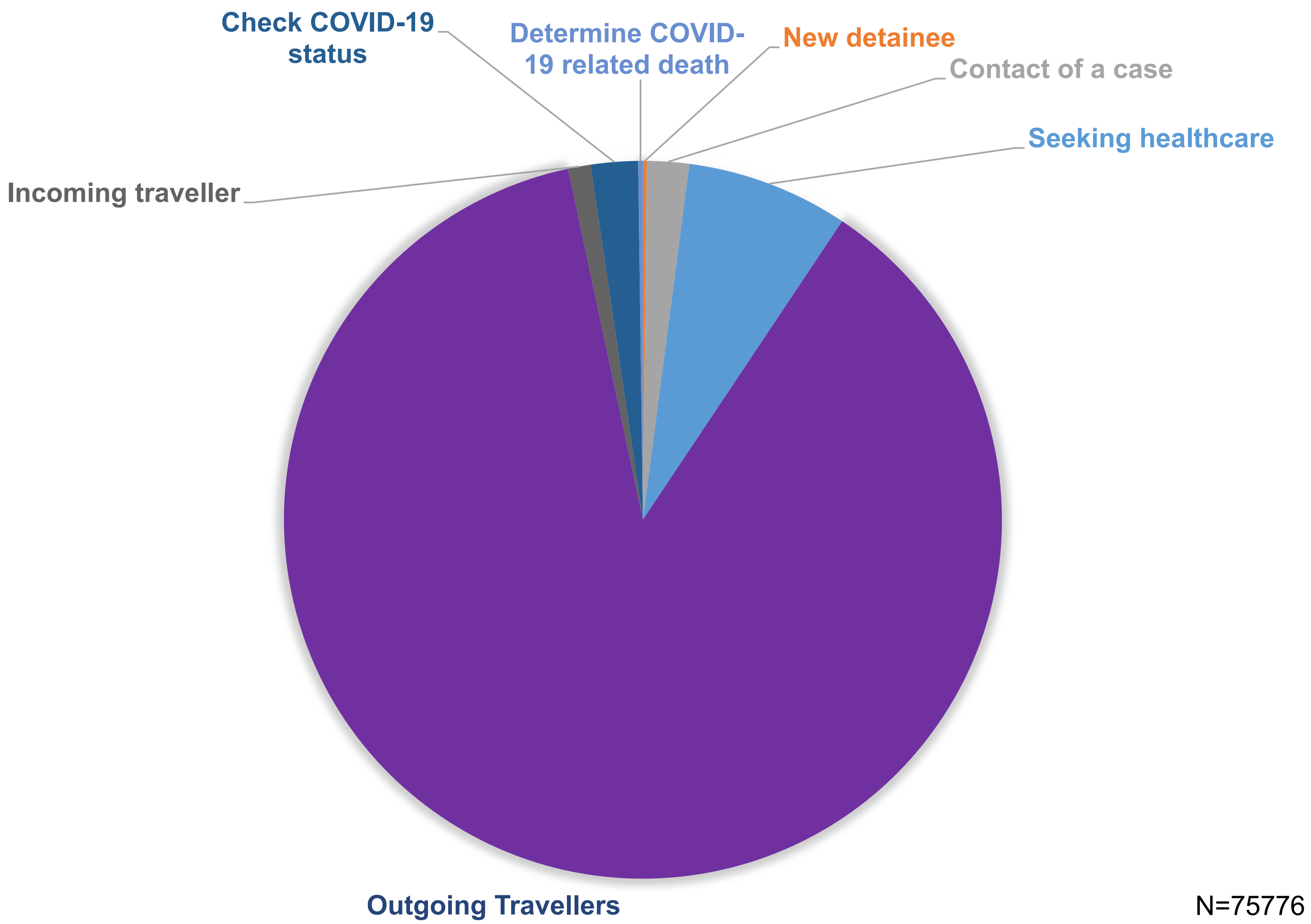

### Supplemental Figure 2

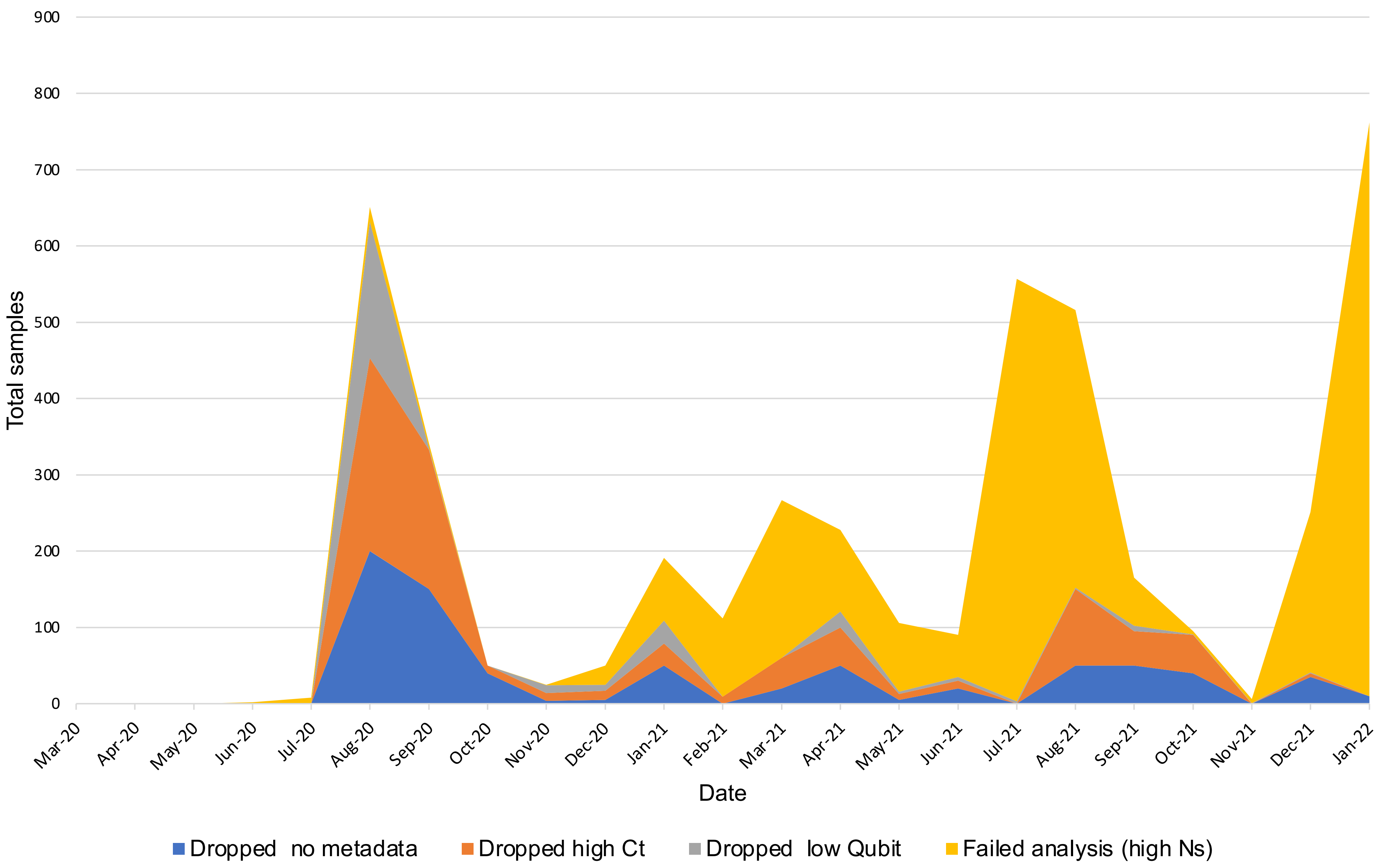

### Supplemental Figure 3

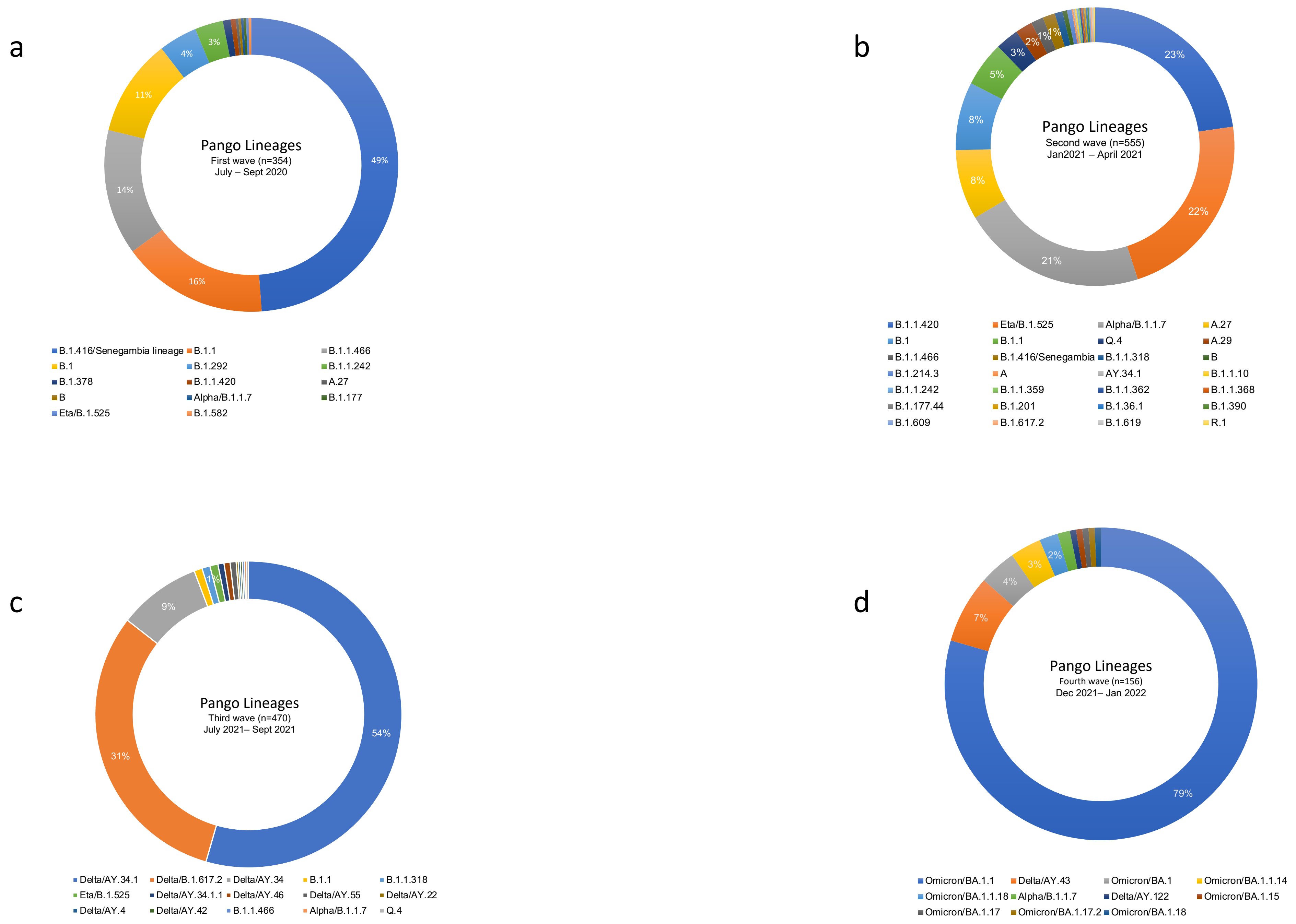
