## Supplementary material for "Genomic epidemiology of SARS-CoV-2 infections in The Gambia, March 2020 to Jan 2022": Ethical approval letter

C/o MRC Unit The Gambia @ LSHTM, Fajara
P.O. Box 273, Banjul
The Gambia, West Africa


**The Gambia Government/MRCG Joint**

### *ETHICS COMMITTEE*

7 September 2022

Dr Abdul Karim Sesay
MRC Unit The Gambia at LSHTM, Fajara

Dear Dr Sesay

**Request ID/Ethics ref:** L2022.E06

**Request Title:** Request for ethics approval waiver

Thank you for submitting your application which was considered by the Gambia Government/MRCG Joint Ethics Committee.

**The documents reviewed were:**

| Document Type/Name | Version Number | Date |
|---|---|---|
| Request letter | | 16 August 2022 |
| Manuscript: Genomic epidemiology of SARS-CoV-2 Infections in The Gambia | | |

On behalf of the committee, I hereby confirm that the request for ethical approval waiver is granted for this manuscript.

With best wishes

Yours sincerely

MK Cham

**Dr Mohammadou Kabir Cham**
**Chairperson, Gambia Government/MRCG Joint Ethics Committee**
